# Hymecromone: A Clinical Prescription Hyaluronan Inhibitor for Efficiently Blocking COVID-19 Progression

**DOI:** 10.1101/2021.10.19.21263786

**Authors:** Shuai Yang, Yun Ling, Fang Zhao, Wei Li, Zhigang Song, Lu Wang, Qiuting Li, Mengxing Liu, Ying Tong, Lu Chen, Daoping Ru, Tongsheng Zhang, Kaicheng Zhou, Baolong Zhang, Peng Xu, Zhicong Yang, Wenxuan Li, Yuanlin Song, Jianqing Xu, Tongyu Zhu, Fei Shan, Wenqiang Yu, Hongzhou Lu

**Affiliations:** Laboratory of RNA Epigenetics, Institutes of Biomedical Sciences & Shanghai Public Health Clinical Center & Department of General Surgery, Huashan Hospital, Cancer Metastasis Institute Shanghai Medical College, Fudan University, Shanghai, China; Shanghai Public Health Clinical Center, Fudan University, Shanghai, China; Zhongshan Hospital, Fudan University, Shanghai, China

## Abstract

**Background:** We previously found that human identical sequences (HIS) of SARS-CoV-2 promote the clinical progression of COVID-19 by upregulating hyaluronan (HA). As one of the drugs for hyaluronan inhibition, hymecromone was chosen for evaluating its therapeutic effects on COVID-19.

**Methods:** ELISA was performed to detect the level of HA in COVID-19 patients. We first analyzed the correlation between the level of plasma HA and clinical parameters (lymphocytes, C-reactive protein, D-dimer, and fibrinogen). We then assessed the correlation between the plasma HA level and pulmonary lesions, which were quantified by using artificial intelligence based on chest CT scans, including ground-glass opacity (GGO) and consolidation. Furthermore, we assessed the effect of hyaluronan treatment on the formation of pulmonary lesions in mice and evaluated the role of hymecromone on hyaluronan production in cultured cells. Finally, 94 of the 144 confirmed COVID-19 patients received oral hymecromone in addition to standard care, whereas the others with only standard care were treated as control. Abnormal serological markers in two groups were selected to determine the efficacy of hymecromone.

**Findings:** Plasma HA was closely relevant to clinical parameters, including lymphocytes (n = 158; *r* = -0.50; *P* < 0.0001), CRP (n = 156; *r* = 0.55; *P* < 0.0001), D-dimer (n = 154; *r* = 0.38; *P* < 0.0001), and fibrinogen (n = 152; *r* = 0.37; *P* < 0.0001), as well as the mass (n = 120; *r* = 0.30; *P* = 0.0008) and volume (n = 120; *r* = 0.30; *P* = 0.0009) of GGO, the mass (n = 120; *r* = 0.34; *P* = 0.0002) and volume (n = 120; *r* = 0.35; *P* < 0.0001) of consolidation. Mice experiment further verified that hyaluronan could cause pulmonary lesions directly. Hymecromone remarkably reduced HA via downregulating *HAS2/HAS3* expression. Accordingly, the number of lymphocytes recovered more quickly as the fold change of lymphocytes per day was higher in hymecromone-treated patients (n = 8) than the control group (n = 5) (*P* < 0.01). Moreover, 89% patients with hymecromone treatment had pulmonary lesion absorption while only 42% patients in control group had pulmonary lesion absorption (*P* < 0.0001).

**Interpretation:** Hyaluronan is closely correlated with COVID-19 progression and can serve as a plasma biomarker. As a promising treatment for COVID-19, hymecromone deserves our further efforts to determine its effect in a larger cohort of COVID-19 patients.

**Funding:** National Key R&D Program of China, Major Special Projects of Basic Research of Shanghai Science and Technology Commission, and Shanghai Science and Technology Innovation Action Plan, Medical Innovation Research Special Project, Research of early identification and warning of acute respiratory infectious diseases.

**Research in context:** *Evidence before this study:* Our previous study revealed that human identical sequences (HIS) of SARS-CoV-2 promotes hyaluronan production in COVID-19 patients. We searched PubMed for studies associated with hyaluronan and COVID-19 using the search terms (“hyaluronan” OR “hyaluronic acid” OR “hymecromone”) AND (“COVID-19” OR “SARS-CoV-2”) without any language restrictions from inception up to May 27, 2021. The studies showed that hyaluronan was present in lung alveoli of severe COVID-19 and SARS-CoV-2 infection-induced hyaluronan. Meanwhile, one report showed that hyaluronan was related to the severity of COVID-19 based on the research of 32 COVID-19 cases. As the inhibitor of hyaluronan synthesis, hymecromone is already an approved drug for patients with biliary spasms in Europe and Asia. However, it is unclear whether hymecromone is an effective therapeutic drug for COVID-19.

*Added value of this study:* We found significant correlations between hyaluronan and clinical parameters (lymphocytes, C-reaction protein, D-dimer, fibrinogen, and pulmonary lesions) in COVID-19 patients. Hyaluronan is the essential material for the induction of ground-glass opacity formation in the lung of COVID-19 patients. The lymphopenia of COVID-19 may be due to T cell exhaustion caused by hyaluronan. Notably, we demonstrated that hymecromone could accelerate the recovery of lymphopenia and pulmonary lesion absorption of COVID-19 in clinical sets.

*Implications of all the available evidence:* Our finding shows that hymecromone could significantly improve the clinical manifestations, especially in severe COVID-19 patients. Reducing hyaluronan using specific drugs could be a promising and alternative therapeutic strategy for COVID-19, especially for the treatment of patients with lymphopenia and pulmonary lesion.x

## Introduction

As of September 17th 2021, more than 226 million infections of COVID-19 have been confirmed worldwide, resulting in approximately 4,666,334 deaths according to the WHO Coronavirus Disease (COVID-19) Dashboard. Significantly, the emergence of B.1.617.2 (Delta) variant has caused breakthrough infections and exacerbated this epidemic^1^, which highlights the importance of exploring the common strategy for the treatment of COVID-19 caused by diverse SARS-CoV-2 variants. With such a rapid variation and high mortality of SARS-CoV-2, there is an urgent need for appropriate treatment to prevent the deterioration of moderate and severe COVID-19, and to reduce the mortality.

Recently, we identified five identical sequences between the genomes of SARS-CoV-2 and human, termed human identical sequences (HIS), which can promote the accumulation of hyaluronan by activating hyaluronic acid synthase 2 (HAS2)^2^. Accordingly, adult respiratory distress syndrome (ARDS) is one of the typical clinical symptoms in severe COVID-19 patients^3^, and hyaluronan (hyaluronic acid, HA) is accumulated in the lung of patients with ARDS^4^. Of note, hyaluronan is higher in lung tissue of deceased COVID-19 patients than that in healthy people^5^. Hyaluronan regulates diverse biological and pathological processes involved in inflammation responses, immune responses, and tissue injury^6^. Meanwhile, the other common features of severe COVID-19 patients include inflammatory cytokine storm, lymphocytopenia, and ground-glass opacity (GGO) in the lung^7,8^. However, the alteration of hyaluronan and its potential role is still unclear.

Here, we found hyaluronan was markedly increased in patients with pulmonary lesions and that there is a significant correlation between plasma level of hyaluronan and other clinical parameters in COVID-19 patients, including lymphocytes, C-reactive protein, D-dimer, and fibrinogen. Moreover, we observed that hyaluronan was significantly relevant to pulmonary lesions, including GGO and the consolidation of lung in COVID-19 patients, which further confirmed that hyaluronan directly induced GGO and the consolidation of lung in mice. Notably, we aimed to use the inhibitor of HA synthesis, hymecromone, an approved prescription drug used for treating biliary spasm in Europe and Asia^9^, to evaluate its therapeutic effect for COVID-19.

## Materials and Methods

### Patient enrollment and experiment animal

All COVID-19 patients enrolled in this study have received a written informed consent upon admission into the Shanghai Public Health Clinical Center (SPHCC), the designated hospital for COVID-19 patients in Shanghai, China^10^. This study was approved by the Ethics Committee of the SPHCC (YJ-2020-S123-02). COVID-19 for patients was confirmed based on the Guidelines of the Diagnosis and Treatment of New Coronavirus Pneumonia (version 7) published by the National Health Commission of China. Exclusion criteria for COVID-19 patients were listed as below: 1) Serious non-infectious pulmonary diseases, including pulmonary tumor, pulmonary edema, atelectasis, pulmonary embolism, pulmonary eosinophilic infiltration, pulmonary vasculitis etc.; 2) Severe liver and kidney dysfunction: a) ALT and AST value were more than 10 times higher than the upper limit of normal value; b) serum creatinine value was more than 1.5 times higher than the upper limit of normal value; c) total bilirubin was more than 2 times the upper limit of normal value; 3) Patients with biliary obstruction; 4) Pregnant women (urine or serum pregnancy test positive) or lactating women; 5) Other factors considered unsuitable by the researchers for this trial, or the situation that may increase the risk of subjects or interfere with the clinical trial.

Adult C57BL/6 mice were purchased from Shanghai Jiesijie experimental animal Co., Ltd (Shanghai, China). All mice were 6–8 weeks of age. Handling of animals was conducted in accordance with the Guide for the Care and Use of Laboratory Animals and were approved by the SPHCC Ethics Committee.

### Procedures

Laboratory parameters, chest computed tomographic (CT) scans, treatment and outcome data were collected according to the patients’ medical records. The mild and severe cases of COVID-19 were distinguished by pulmonary lesions based on chest CT. The mild COVID-19 patients did not have pulmonary lesions, while the severe COVID-19 patients had pulmonary lesions including GGO and/or consolidation. We first detected the plasma hyaluronan levels of COVID-19 patients (n=158) in SPHCC using Enzyme-linked Immunol sorbent assay (ELISA) as described previously^2^. Meanwhile, twenty health subjects were recruited to evaluate their plasma hyaluronan levels as the control group. The 48.43 ng/ml of hyaluronan was sensitive to distinguish COVID-19 patients and health subjects via receiver operating characteristic curve (ROC) analysis. Then, we divided the COVID-19 patients into two groups (HA ≥ 48.43 ng/ml; HA < 48.43 ng/ml) and confirmed whether there are significant differences between hyaluronan and other clinical parameters (lymphocytes, C-reactive protein, D-dimer, and fibrinogen). The lesion regions in the lungs of COVID-19 patients were quantified by artificial intelligence (AI) as previous description^11,12^. Furthermore, we analyzed the correlation between hyaluronan and these clinical parameters.

Adult male mice were used to assess the impact of hyaluronan on the lung lesions, such as GGO and consolidation. The 200 to 400 kDa of hyaluronan dissolved in 1×PBS was intratracheal to mice (60 mg/kg), while the 1×PBS treatment was as the control group. Then, the formation of lung lesions in two groups were monitored via QuantumGX microCT on day four.

Moreover, we evaluated whether hymecromone can reduce hyaluronan levels in the cell using HEK293T and HUVEC cells. Twenty-four hours after HEK293T and HUVEC were treated with DMSO or hymecromone (250 μg/ml), cell culture mediums were collected to detect the hyaluronan levels using the Hyaluronan DuoSet ELISA (R&D Systems) according to the manufacturer’s descriptions. Meanwhile, we extracted the total RNA of HEK293T and HUVEC cells treated with DMSO or hymecromone. As previously described in our study^2^, quantitative RT-PCR (RT-qPCR) was performed to evaluate whether hymecromone can decrease the expression of *HAS1, HAS2*, and *HAS3* (*HAS1/2/3*), which are the known hyaluronic acid synthases. The expression of *GAPDH* served as the normalized endogenous control. Relative mRNA expression was calculated via the 2^-ΔΔCt^ method (The primers are shown in Supplementary Table 1).

Finally, 144 COVID-19 patients were recruited to assess whether hymecromone could improve the clinical parameters of COVID-19. Among these patients, 94 patients were oral hymecromone administration (2 tablets, 0.2 g/tablet, three times a day, before meals) combined with conventional treatment as the experimental group. The other 50 patients only underwent conventional treatment as the control group. Administration of hymecromone continued until the patients recovered from COVID-19. Chest CT scans, lymphocyte counts, C-reactive protein (CRP), D-dimer, fibrinogen, and plasma hyaluronan were critical clinical indicators during COVID-19 patient treatment.

### Outcomes

Outcomes of COVID-19 patients were assessed based on the clinical indicators and chest CT images. The primary outcomes were the changes in lymphocyte counts, CRP, fibrinogen, and D-dimer in patients. The secondary outcome was the change in the patients’ chest CT results. We also monitored the suspected serious adverse reactions in accordance with regulatory requirements.

### Statistical analysis

The COVID-19 patients with pulmonary lesions were defined as severe while the others were considered mild. Twenty health subjects without any clinical symptoms were defined as the normal group. Samples (n=158) were collected to compare the levels of hyaluronan in these three groups using the Mann–Whitney test. ROC analysis was used to identify the hyaluronan concentration for distinguishing normal groups and COVID-19 patients. Based on this analysis, COVID-19 patients were divided into two groups (HA ≥ 48.43 ng/ml; HA < 48.43 ng/ml). The significant differences between typical clinical indicators (such as lymphocytes, C-reactive protein, D-dimer, and fibrinogen) in these two groups were calculated by the Mann–Whitney test. Two-tailed Spearman’s correlation analysis was performed to evaluate the correlation between hyaluronan and these clinical indicators. Unpaired T test was used to confirm the significant differences between hyaluronan and *HAS1/2/3* expression among hymecromone-treated cells and DMSO-treated cells.

In addition, 144 COVID-19 patients were recruited to assess the effect of hymecromone for COVID-19. We further analyzed the effect of hymecromone using these patients with abnormal clinical parameters, including lymphocytes, C-reactive protein, D-dimer, and fibrinogen. For the primary outcomes, we calculated the fold changes of these clinical parameters per day in the hymecromone-treated group and control group and compared the clinical significances using the Mann–Whitney test. For the secondary outcome of change in chest CT results, we compared the hymecromone-treatment group and the control group, including the patients with GGO. According to CT quantitative analysis, we evaluated the change of pulmonary lesions, including improvement and exacerbation in COVID-19 patients classified by different hospitalization days (X < 14 day; 14 day ≤ X < 28 day; 28 day ≤ X < 35 day; X represents the hospitalization days). We calculated the percentage of improvement and exacerbation in different groups and analyzed the significance between the hymecromone-treated group and the control group using Fisher’s exact tests. The *P* value less than 0.05 was statistically significant (*, *P* < 0.05; **, *P* < 0.01; ***, *P* < 0.001; ****, *P* < 0.0001; ns, not significant).

## Results

### Hyaluronan is a considerable biomarker to predict COVID-19 progression

A total of 158 COVID-19 patients in SPHCC were conducted to investigate the potential relationship between hyaluronan and the typical clinical indicators for COVID-19. Specifically, 18% (28 of 158) patients without pulmonary lesions were mild, while the other patients (130 of 158) with pulmonary lesions including GGO and consolidation were severe. The plasma HA level showed no significant difference between healthy subjects and mild COVID-19 patients, while the plasma HA level in severe COVID-19 patients were significantly higher than that in the other two groups (figure 1A), indicating there is a potential relation between hyaluronan and pulmonary lesions. Further ROC analysis identified that the 48.43 ng/ml of hyaluronan was sensitive for distinguishing COVID-19 patients and healthy subjects (figure 1B). Then, 158 COVID-19 patients were divided into low HA group (HA <48.43 ng/ml) and high HA group (HA ≥ 48.43 ng/ml).

**Figure 1:**
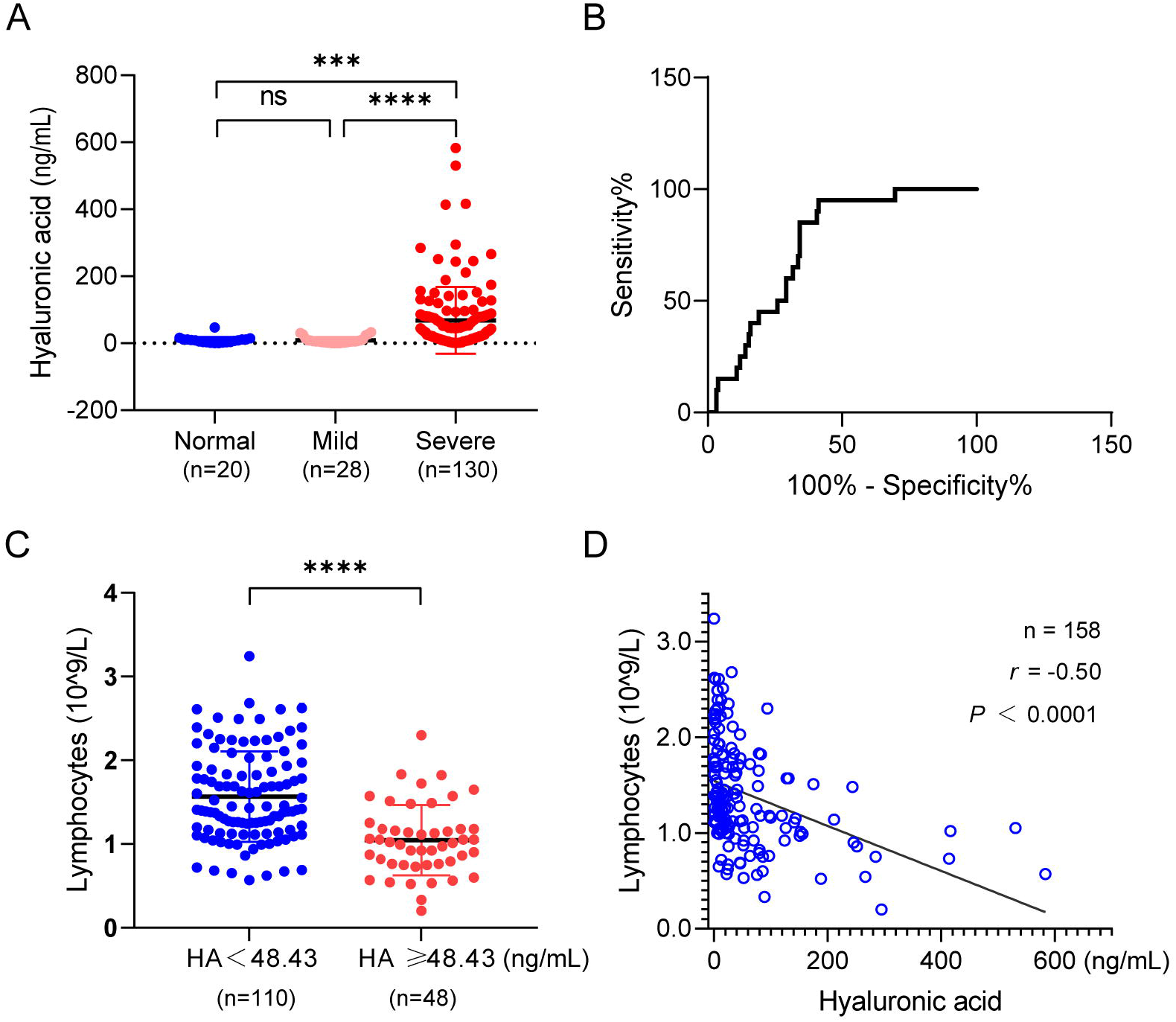
The increase of hyaluronan is accompanied with the severity in COVID-19 patients. (A) The plasma hyaluronan level of COVID-19 patients and normal health subjects were evaluated by ELISA. The classification of mild and severe of COVID-19 was based on ground-glass opacity (GGO) of chest CT. (B) ROC of the plasma hyaluronan level in normal volunteers and COVID-19 patients. The 48.43 ng/ml of hyaluronan is sensitivity value to distinguish the normal subjects and COVID-19 patients. (D) Counts of lymphocytes upon admission plotted against hyaluronic acid. Two-tailed Spearman’s correlation analysis was performed to evaluate the correlation between hyaluronan and lymphocyte counts. The significant difference in (A) and (C) was analyzed by the Mann–Whitney test. *, *P* < 0.05; **, *P* < 0.01; ***, *P* < 0.001; ****, *P* < 0.0001; ns, not significant.

Lymphopenia is one of the typical clinical symptoms in severe COVID-19 patients^13^. It is reported that hyaluronan can result in the death of the activated T cell ^14^. Here, we found that lymphocytes were markedly decreased in the high HA group of COVID-19 patients (figure 1C). As shown in figure 1D, hyaluronan was negatively correlated with lymphocytes (n = 158; *r* = -0.50; *P* < 0.0001). The subsets of T lymphocytes, CD4+ T cells, CD8+ T cells, and CD45+ T cells were also significantly reduced in COVID-19 patients with high HA (supplemental figure 1A-C). Similarly, hyaluronan was negatively correlated with the CD4+ T cells (n = 151; *r* = -0.44; *P* < 0.0001), CD8+ T cells (n = 151; *r* = -0.53; *P* < 0.0001), and CD45+ T cells (n = 151; *r* = -0.49; *P* < 0.0001) (supplemental figures 1D-F).

Inflammation is another common clinical symptom involved in COVID-19 patients, usually assessed via CRP. Noteworthily, low molecular weight of hyaluronan is an important inflammation mediator.^6^ Surprisingly, we found that CRP markedly increased in the high HA group (figure 2A) and was positively correlated with hyaluronan (n = 156; *r* = 0.55; *P* < 0.0001) (figure 2B). Recent studies revealed abnormal blood coagulation in COVID-19 patients^7^. Clinically, D-dimer and fibrinogen are commonly used to assess a patient’s blood coagulation. We found that both were higher in COVID-19 patients with high HA and were positively correlated with hyaluronan (figures 2C-F).

**Figure 2:**
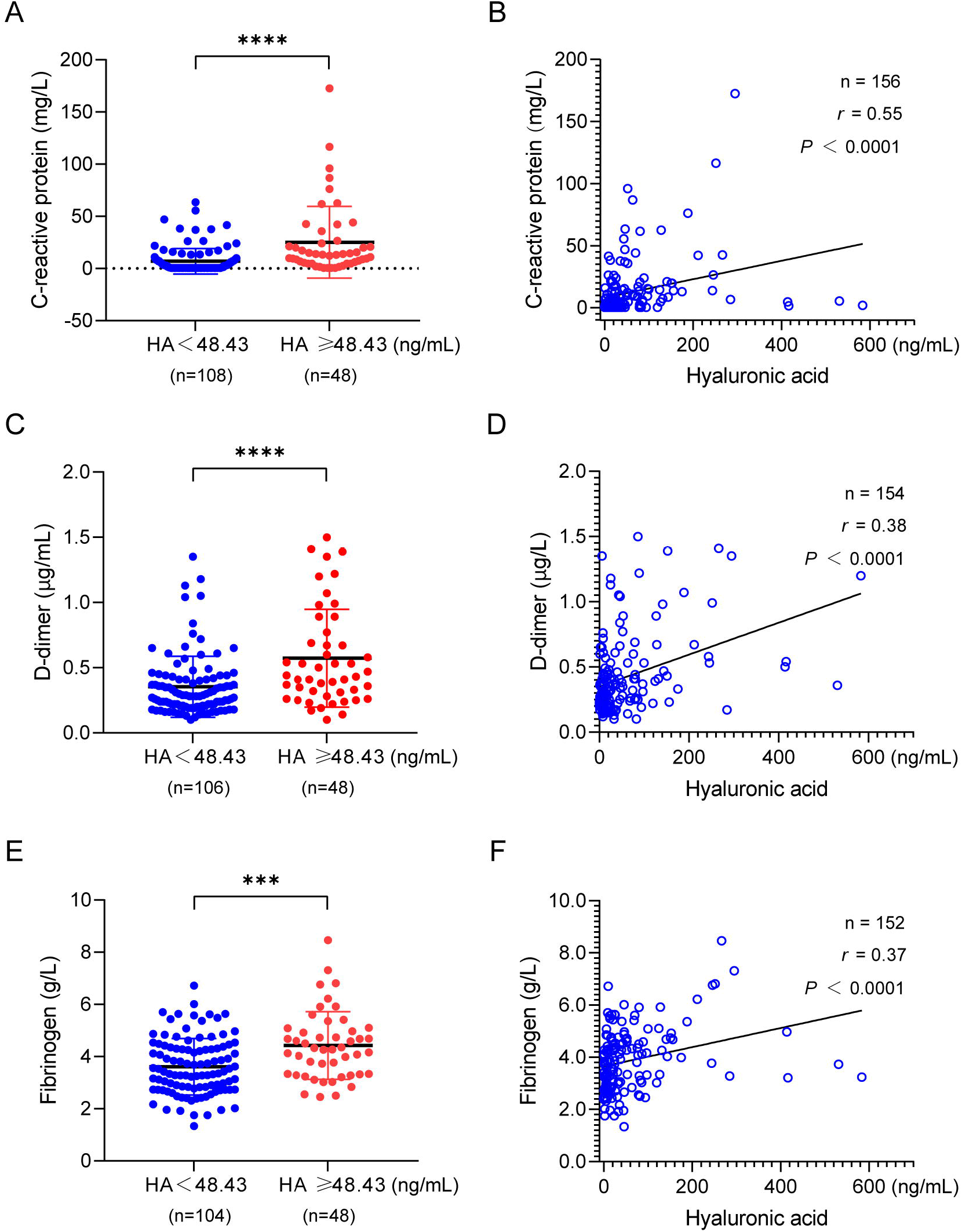
The plasma level of hyaluronan as a typical indicator of COVID-19 patients in clinic. C-reactive protein (A), D-dimer (C), and fibrinogen (E) of COVID-19 patients were showing based on the 48.43 ng/ml of hyaluronan. Data was presented by Mean ± SD. The significant difference was confirmed by the Mann–Whitney test. *, *P* < 0.05; **, *P* < 0.01; ***, *P* < 0.001; ****, *P* < 0.0001; ns, not significant. The relation of C-reactive protein (B), D-dimer (D), and fibrinogen (F) against hyaluronic acid were determined by two-tailed Spearman’s correlation analysis.

Collectively, these results reveal that the increase of hyaluronan is significantly relevant to the reduction of lymphocytes and the upregulation of CRP, D-dimer, and fibrinogen in COVID-19 patients, suggesting hyaluronan may play a key role during the clinical progression of COVID-19.

### Hyaluronan is fundamental for ground-glass opacity formation in the lung of COVID-19 patients

It is well-known that ground-glass opacity of lungs is the other typical clinical manifestation of COVID-19 patients^15^, which can develop into consolidation. We quantified the mass and volume of the lung lesion regions involved in GGO and consolidation in 120 COVID-19 patients using uAI-Discover-NCP (beta version). In general, GGO was defined in a range from -750 HU to -300 HU, and the consolidation region was defined from -300 HU to 50 HU^16^. Notably, hyaluronan was positively correlated with the mass (n = 120; *r* = 0.30; *P* = 0.0008) and volume (n = 120; *r* = 0.30; *P* = 0.0009) of GGO, which was also the case in consolidation (supplemental figure 2). A typical case for this correlation is shown in figure 3. The plasma HA levels, GGO, and consolidation clearly increased on the fourth day, compared to the first day (figures 3A-C), indicating the mass and volume of pulmonary lesions (GGO and consolidation) increased along with the upregulation of hyaluronan. CT images further showed the exacerbation of pulmonary lesions, including GGO and consolidation in this patient on the fourth day instead of the first day (figure 3D). Recent research found that there is jelly-like liquid in the lung of COVID-19 patients^3^. Given that hyaluronan can absorb water reaching 1000 times its molecular weight^17^, we hypothesize that hyaluronan may be one of the determinants for GGO of lung in COVID-19 patients.

**Figure 3:**
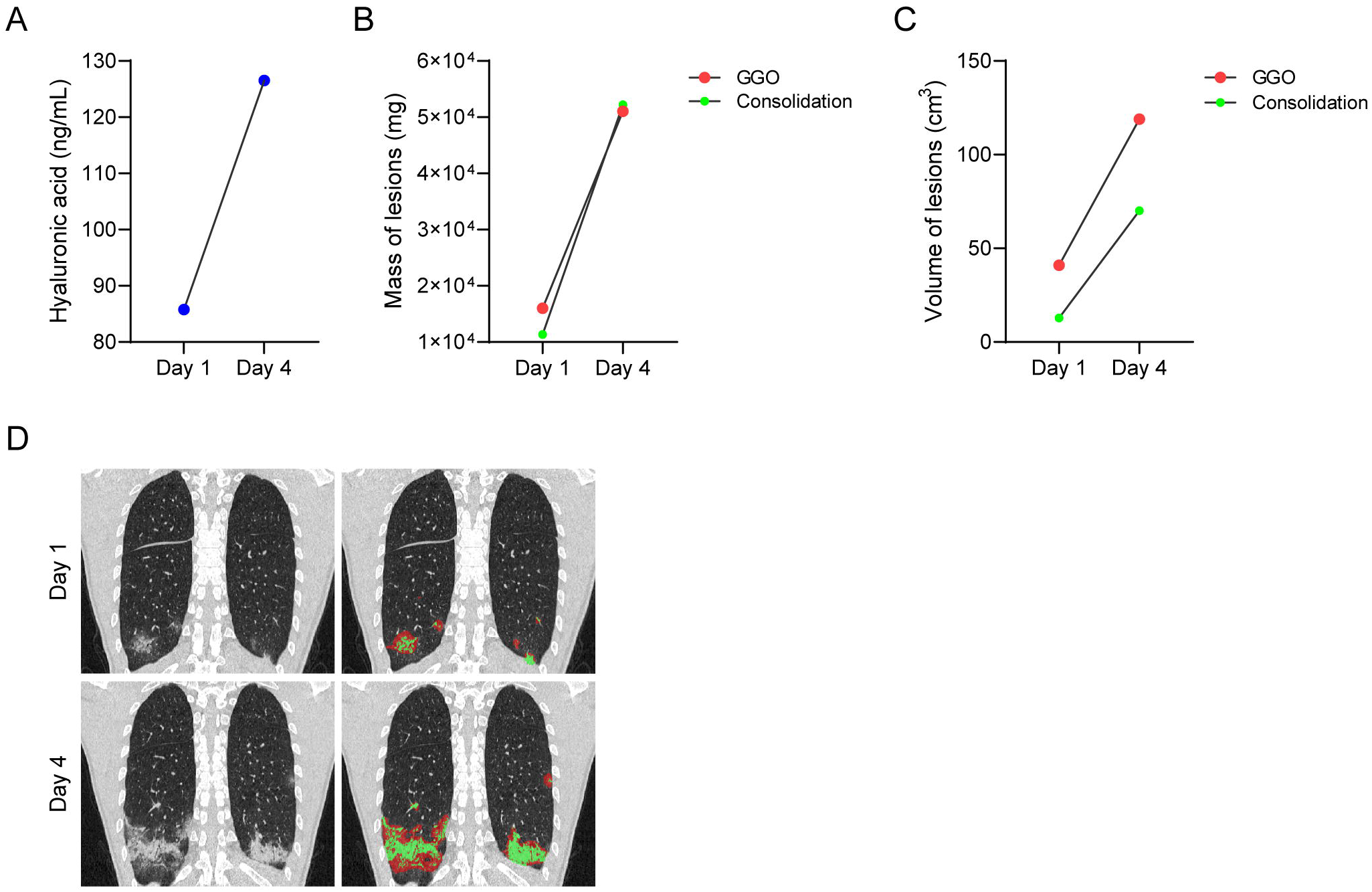
The size of pulmonary lesions and the level of hyaluronan in one typical case. (A) The plasma HA levels of this case were detected at the first day and fourth day. (B-C) The mass (B) and volume (C) of pulmonary lesions including GGO and consolidation were calculated via the automatic lung segmentation technology of AI based on CT images of this case at the first day and fourth day. (D) Represented CT images showing the pulmonary lesions in this case. The original CT results were shown in black-and-white images while the marked lesion regions of CT results were shown in color images. Red regions indicated GGO, and green regions indicated consolidation.

To confirm our hypothesis, we delivered hyaluronan intratracheally to the lung in male mice. Significantly, CT images showed that GGO and consolidation of lung occurred in mice treated with hyaluronan while there were no pulmonary lesions in mice treated with 1 × PBS (figure 4). Therefore, these findings supported that hyaluronan acts as a critical material for the formation of GGO and consolidation of the lung in COVID-19 patients, indicating that inhibition of HA synthesis may be a promising strategy for relieving pulmonary lesions in COVID-19 patients.

**Figure 4:**
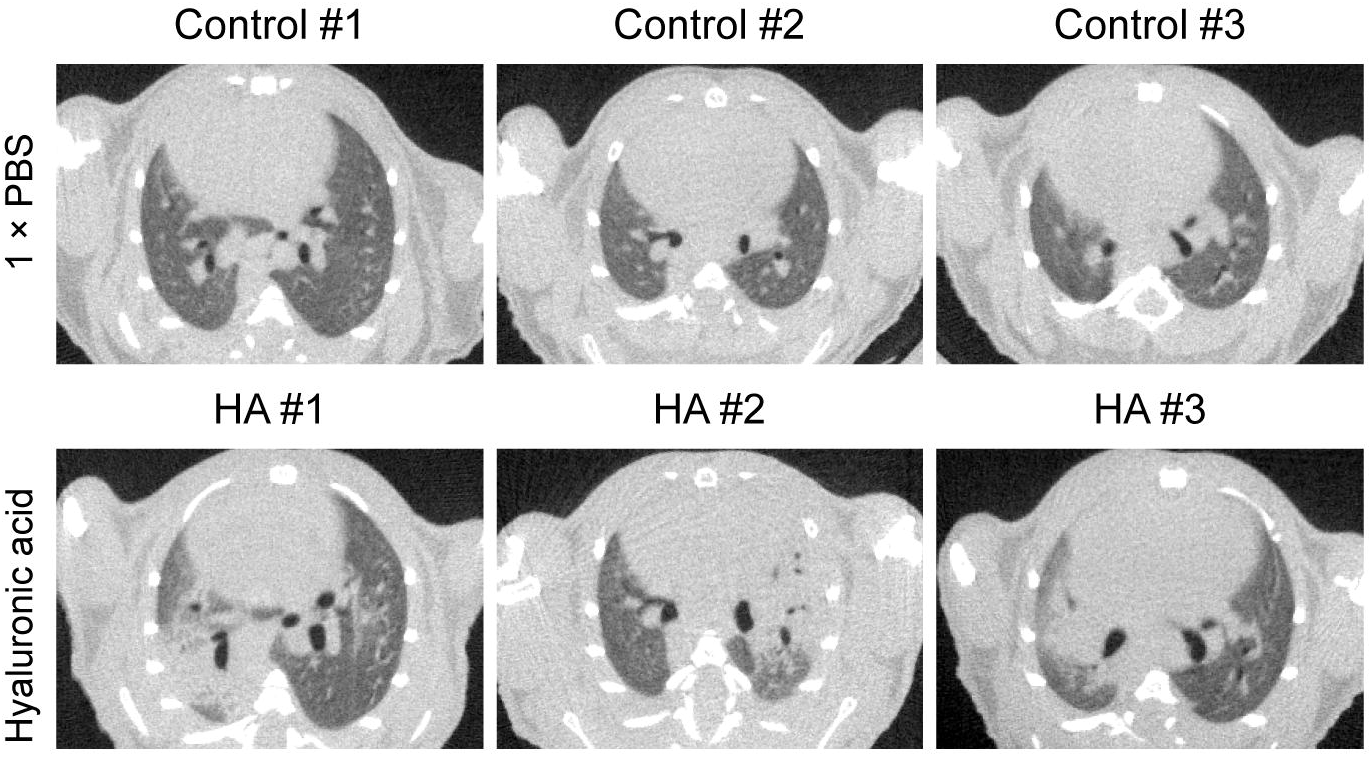
Hyaluronan directly induces GGO and consolidation in adult mice. Represented CT images of lungs in mice with different treatment were shown. Adult C57BL/6 mice were used to assess whether hyaluronic acid induces pulmonary lesions. We directly delivered hyaluronan (200∼400 kDa) to the trachea (n = 3), and 1×PBS treatment was as the control group (n = 3). Then, we monitored the lungs of mice in two groups via QuantumGX microCT at the fourth day.

### Hymecromone significantly decreases hyaluronan bydownregulating *HAS2/HA3*

Based on these findings, we realized that reduction of HA production could be an alternative therapeutic strategy for COVID-19, especially for patients with pulmonary lesions. As a derivative of coumarin, 4-MU was shown to inhibit the production of HA^18^. Fortunately, we noticed there is a commercial drug, 4-MU, also called hymecromone, which is an approved prescription drugs in China, USA and Europe, even acting as over the counter drugs in some areas. We first verified the inhibitory effect of hymecromone on HA production in HEK293T and HUVEC cells. As expected, HA from culture medium in HEK293T and HUVEC cells treated with hymecromone (250 μg/ml) were significantly lower than that in DMSO-treated cells (figure 5A). There are three known hyaluronic acid synthases, including *HAS1, HAS2*, and *HAS3* (*HAS1/2/3*). As shown in figures 5B-C, hymecromone treatment remarkably downregulated the expression of *HAS2/HAS3*, but did not affect the expression of *HAS1* in HEK293T and HUVEC cells (figures 5B-C). Therefore, hymecromone inhibits the production of hyaluronan by decreasing *HAS2/HA3* expression.

**Figure 5:**
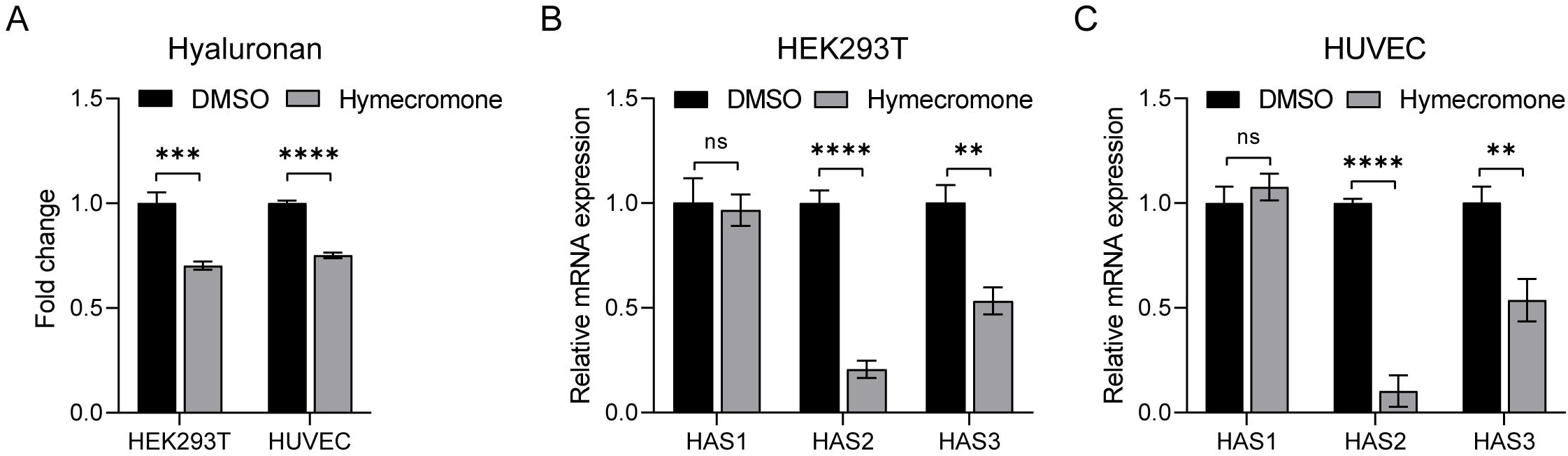
Hymecromone decreases hyaluronan by downregulating hyaluronic acid synthases. (A) ELISA detected the hyaluronic acid of culture medium in HEK293T and HUVEC treated with DMSO or hymecromone (250 μg/ml). The fold change of hyaluronic acid was normalized to DMSO. (B-C) RT-qPCR evaluated the mRNA levels of *HAS1, HAS2*, and *HAS3* in HEK293T (B) and HEK293T treated with DMSO or hymecromone (250 μg/ml). Data was presented by Mean ± SD. The significant difference was confirmed by unpaired t test. *, *P* < 0.05; **, *P* < 0.01; ***, *P* < 0.001; ****, *P* < 0.0001; ns, not significant.

### Hymecromone accelerates the recovery of clinical manifestations in COVID-19 patients

To further assess whether hymecromone is efficient inimproving the clinical parameters of COVID-19, we recruited 144 confirmed COVID-19 patients. Among these patients, 94 (65%) patients with hymecromone treatment were in the clinical trial group, while 50 (35%) patients with support treatment were in the control group (figure 6A). Given the significant correlation between hyaluronan and clinical indicators, we set changes in lymphocytes, CRP, fibrinogen, and D-dimer as the primary endpoints, and the change in chest CT results as the secondary endpoint. To objectively evaluate the effect of hymecromone on COVID-19, we selected COVID-19 patients with abnormal clinical indicators in these two groups. Specifically, there were 5 patients with decreased lymphocytes, 4 patients with increased CRP, 7 patients with increased fibrinogen, and 8 patients with increased D-dimer in the control group. In the clinical trial group, there were 8 patients with decreased lymphocytes, 5 patients with increased CRP, 7 patients with increased fibrinogen, and 6 patients with increased D-dimer (figure 6B).

**Figure 6:**
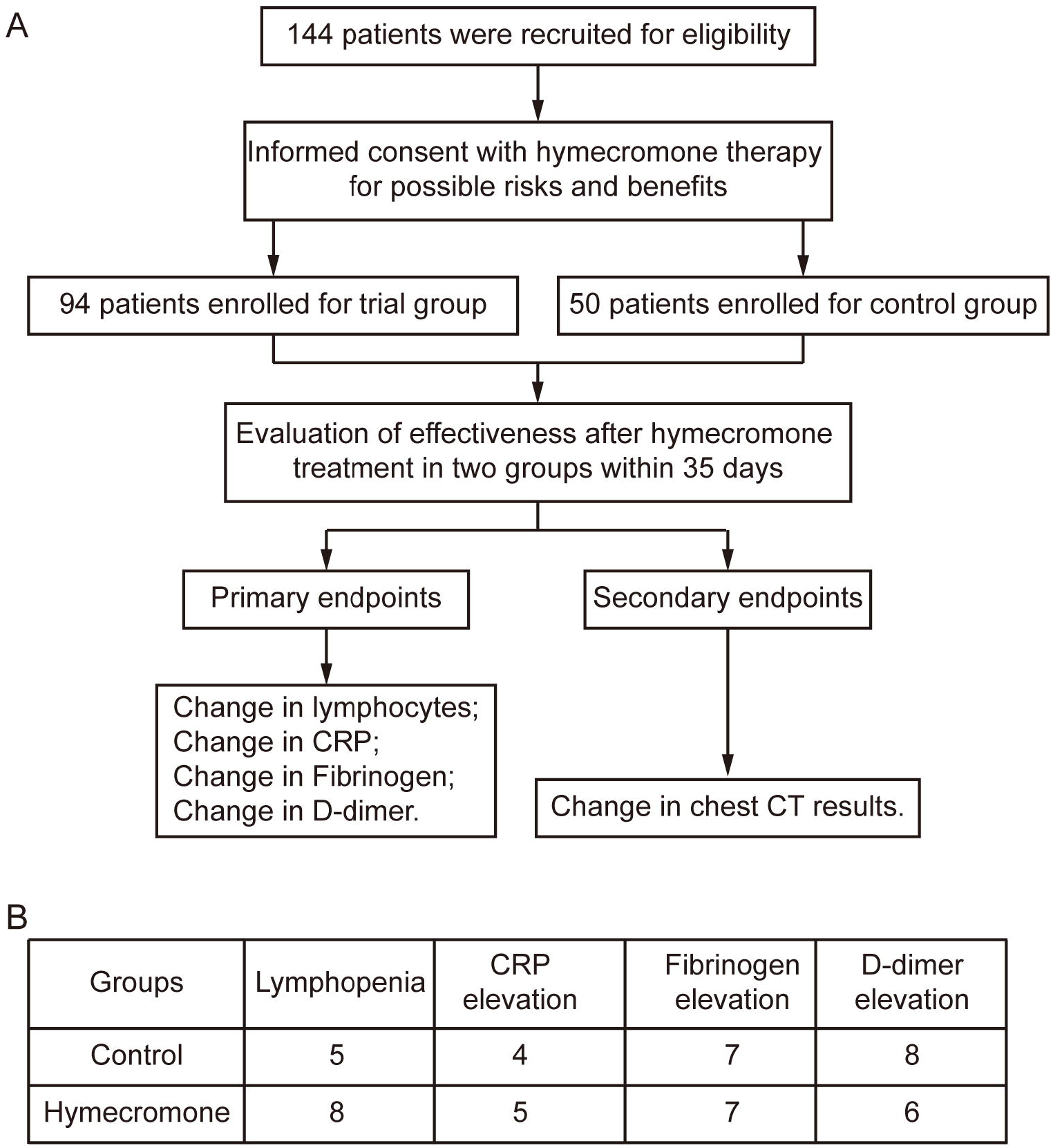
The Consort Diagram and Statistical Table of recruited COVID-19 patients for this clinical trial. (A) The Consort Diagram for the clinical trial contains 144 COVID-19 patients. Among these patients, 94 patients were in the trial group while the rest were in the control group. Changes in lymphocytes, CRP, fibrinogen, and D-dimer were set as the primary endpoints and change in chest CT results were set as the secondary endpoint. (B) Statistical Table of COVID-19 patients with change in clinical indicators of control group and experimental group within 35 days.

Firstly, we focused on the change of pulmonary lesions in COVID-19 patients during hymecromone treatment. We determined the situation of pulmonary lesions, including GGO and consolidation based on CT quantitative analysis. Surprisingly, all the patients with hospitalization days < 14 day had improved pulmonary lesions after hymecromone treatment (figure 7A). The percentage of pulmonary lesion improvement in patients with hymecromone treatment were significantly higher than that in control group (*P* = 0.0002). For the patients with hospitalization between 28 to 35 days, the percentage of pulmonary lesion improvement rate was 86% (6 of 7) patients in the trial group, and 45% (2 of 9) in the control group, respectively. In total, 89% (41 of 46) patients with hymecromone treatment had pulmonary lesion absorption, while only 42% (42 of 100) in the control group had pulmonary lesion absorption (*P* < 0.0001). Different regions of pulmonary lesions in typical cases in two groups were gradually absorbed (figure 7B). As such, patients have better improvement of pulmonary lesions after hymecromone treatment. Thus, hymecromone could promote the pulmonary lesion absorption of COVID-19.

**Figure 7:**
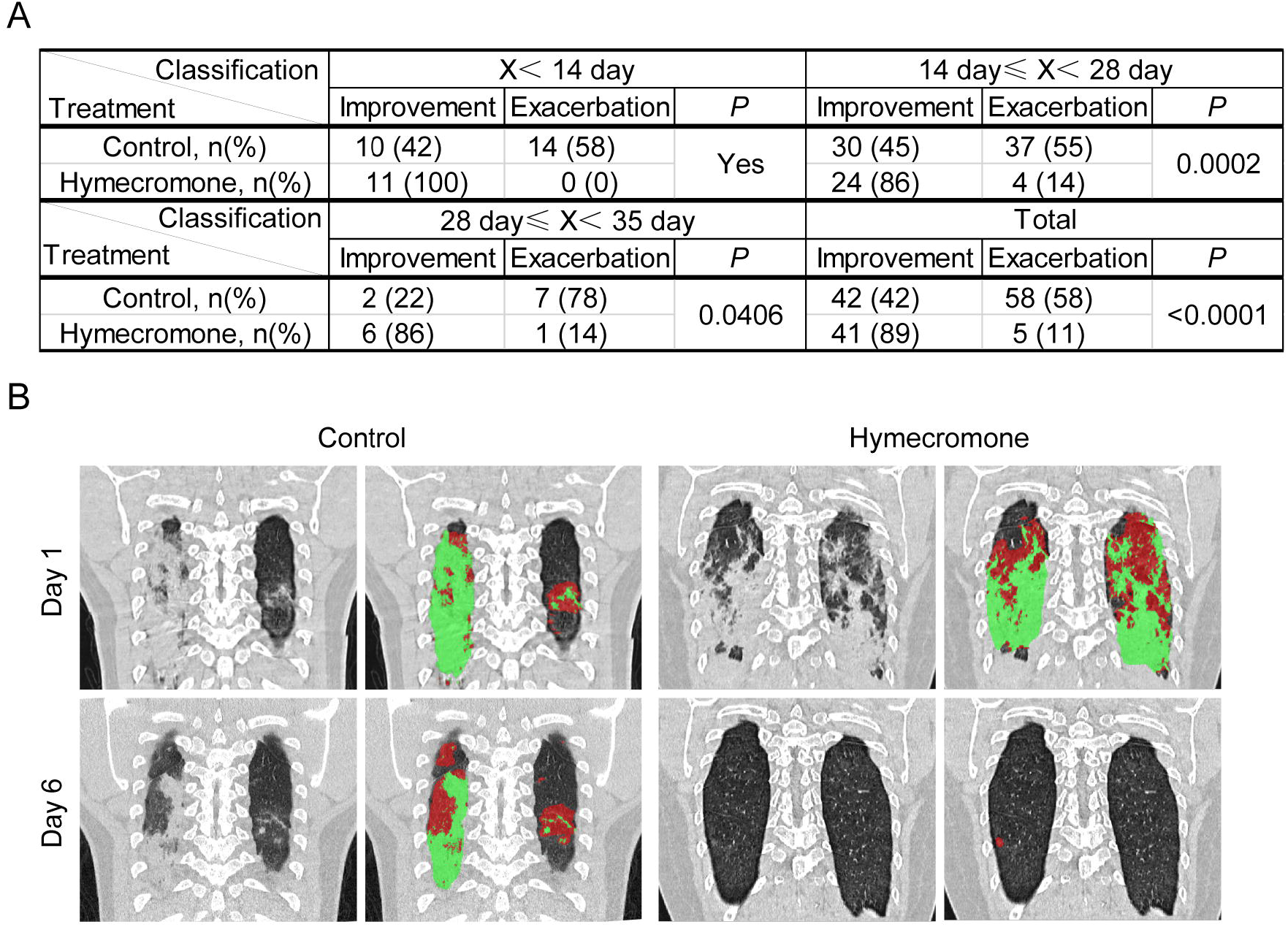
Hymecromone can effectively alleviate the pulmonary lesions of severe COVID-19 patients. (A) Comparison of change in chest CT results between control group and hymecromone treatment group. Fisher’s exact tests were used to evaluate the significance between control group and experiment group. (B) Represented CT images of COVID-19 patients with support or hymecromone treatment. The original CT results were shown in black-and-white images of two groups while the marked lesion regions of CT results were shown in color images of two groups. Red regions indicated GGO, and green regions indicated consolidation.

Then, we calculated and compared the fold changes of these various clinical indicators per day in these two groups with lymphopenia, CRP elevation, fibrinogen elevation, and D-dimer elevation, respectively. As shown in figure 8A, the fold change of lymphocytes per day was remarkably higher in hymecromone-treated patients, compared to the control group. Similarly, the fold changes of CRP or fibrinogen in hymecromone-treated patients were higher than that in the control group (supplemental figures 3-5). These results implied that hymecromone contributes to the improvement of clinical parameters of COVID-19. Moreover, the fold change of D-dimer tended to be improved, but it is not significant between control group and clinical trial group, which may be due to to the limited number of patients.

**Figure 8:**
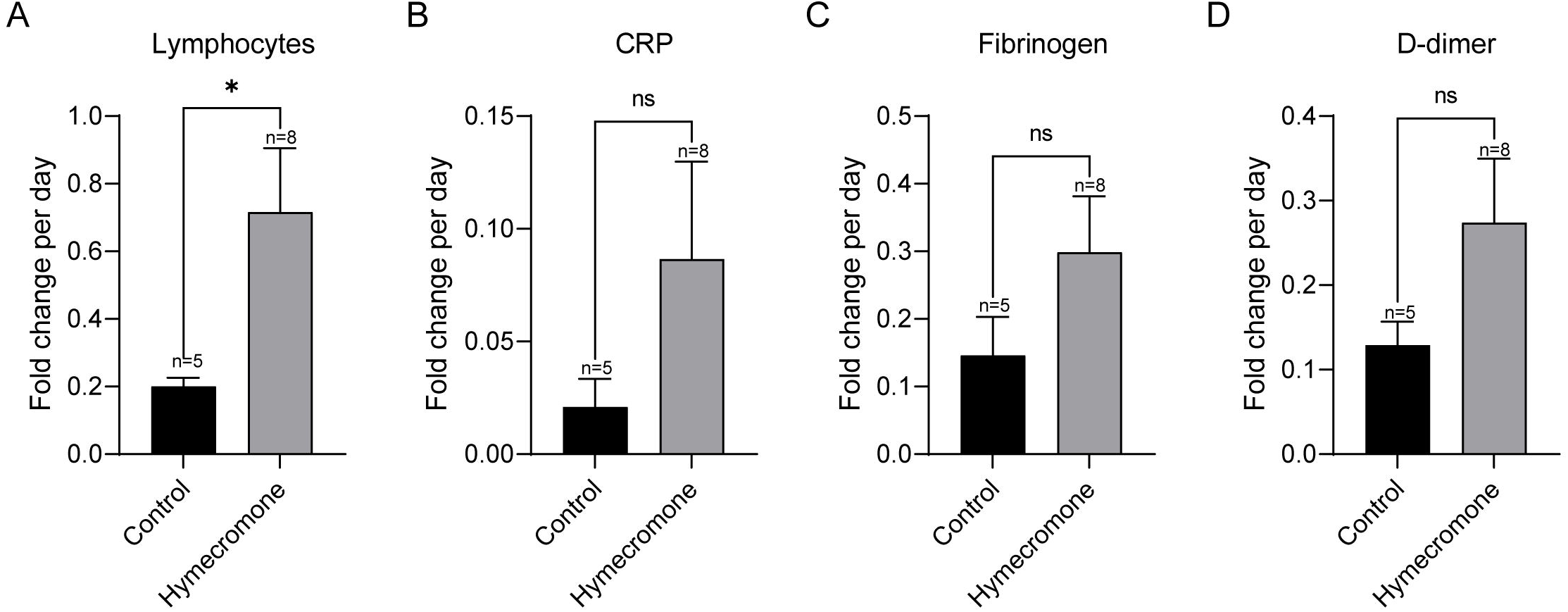
Hymecromone accelerate the recovery of COVID-19 patients from lymphocytopenia. Changes in lymphocytes (A), CRP (B), fibrinogen (C), and D-dimer (D) were calculated as the fold change of diverse clinical indicators per day in patients with lymphocytopenia. Data was presented by Mean ± SD. The significant difference was confirmed by the Mann–Whitney test. *, *P* < 0.05; **, *P* < 0.01; ***, *P* < 0.001; ****, *P* < 0.0001; ns, not significant.

These findings demonstrated that hymecromone is a promising drug for effectively improving the clinical manifestations of COVID-19.

## Discussion

Our previous study has found that human identical sequences (HIS) of SARS-CoV-2 could promote hyaluronan upregulation by NamiRNA-Enhancer network during the progression of COVID-19^2^, indicating that reduction in hyaluronan may be an alternative therapeutic strategy for COVID-19, which is supported by the other reports found that hyaluronan was increased in severe COVID-19 patients^5,19,20^. As an inhibitor of HA synthesis, hymecromone is an approved drug for biliary spasms treatment. Here, we aimed to assess whether hymecromone could promote the prognosis of COVID-19.

Hyaluronan is an excellent biomarker to predict COVID-19 progression. We found that hyaluronan was significantly elevated in severe COVID-19 patients. Consistent with our results, increases in hyaluronan was also confirmed in severe or critical patients with COVID-19^20^. Notably, hyaluronan is accumulated in the bronchoalveolar lavage fluid (BALF) and serum samples of patients with ARDS^21^, a relatively serious COVID-19. We also found that hyaluronan was significantly correlated with lymphocytes, CRP, D-dimer, and fibrinogen. These clinical indicators are proven biomarkers for the clinical progression of COVID-19 ^22^. All these findings demonstrated that hyaluronan was relevant to COVID-19 progression. Additionally, we showed that hyaluronan positively correlates with quantified chest CT results, a recently identified parameter to predict the severity of COVID-19^11^. This result further supported that hyaluronan can serve as a better biomarker for predicting the clinical progression of COVID-19.

Most importantly, our results provide vital insights into the typical clinical symptoms occurring in COVID-19. As we all know, GGO is one of typical CT manifestations of COVID-19 patients^23^. However, the pathophysiologic mechanism of GGO is still unclear. We showed that hyaluronan can cause GGO and consolidation of lung in mice, providing direct evidence that hyaluronan is fundamental for GGO formation. Consistent with our results, transcriptome sequencing of cells in BALF from COVID-19 patients revealed differentially altered genes enriched in the hyaluronan metabolic pathway^19^. The appearance of GGO were further interpreted by the facts that the water absorption of hyaluronan can reach 1000 times its molecular weight^17^, possibly resulting in the formation of jelly-like liquid in the lung of COVID-19 patients^3^. In addition to GGO, lymphopenia is another typical clinical symptom in severe COVID-19 patients^13^. We found that the crucial subsets of T lymphocytes, CD4+ T cells, CD8+ T cells, and CD45+ T cells were also significantly decreased in COVID-19 patients with high HA. Meanwhile, the negative correlation between hyaluronan and diverse T cell subsets confirmed that hyaluronan was associated with the reduction of T lymphocytes. In line with this sight, hyaluronan can induce the activated T cell death by binding to its ligand CD44^14^. Notably, CD4+ T lymphocytes become rapidly activated after SARS-CoV-2 infection^24^, which provides the condition needed for the binding of HA to CD44 located in activated T cells, leading to their death. In other words, T cell exhaustion mediated by hyaluronan may potentially underly lymphopenia of COVID-19 patients.

Hymecromone could accelerate the recovery of clinical manifestations in COVID-19 patients. At present, the therapeutic strategy for COVID-19 is mainly symptomatic supportive treatment in clinic^25^. Here, we found that hymecromone inhibited hyaluronan production by suppressing the expression of *HAS1/2*. Additionally, hymecromone significantly improved lymphopenia of COVID-19 patients, indicating that declines in hyaluronan via hymecromone administration promote the recovery of lymphopenia. Likewise, hymecromone decreased the CRP and fibrinogen elevation of COVID-19 patients. Also remarkably, hymecromone accelerated pulmonary lesions absorption. It has already been reported that dexamethasone and metformin can significantly decrease the mortality of patients with severe COVID-19^26-28^. Accordingly, these results may be explained by the reports that dexamethasone and metformin can also rapidly decline hyaluronan synthesis by downregulating *HAS2* expression^29,30^, which may contribute the therapeutic effects of these two drugs on COVID-19. In addition, we found most HIS is quite conservative by analyzing 159258 genomes of SARS-CoV-2 (unpublished data), suggesting hyaluronan caused by HIS may be an important therapeutic target for diverse SARS-CoV-2 variants. All these evidences support that hymecromone could be a potential and effective drug for COVID-19 patients even infected with delta variant.

There are several limitations in our study. First, the total number of patients involved in our clinical trial is relatively insufficient, which requires a larger sample and multi-center clinical study in the future. Second, we did not evaluate whether hymecromone can reduce the mortality rates of COVID-19 patients because of no critical patients in this clinical trial. Third, given that the exact value of hyaluronan may be different using diverse methods, it is necessary to identify the hyaluronan concentration to clinically distinguish the healthy subjects and COVID-19 patients by uniform standard methods.

Overall, the increased hyaluronan is significantly correlated with the decreased lymphocytes and pulmonary lesions of COVID-19 patients. Hymecromone administration can markedly improve the clinical manifestations of COVID-19 patients. Thus, hymecromone could be a potential and efficient drug for COVID-19 therapy, which is needed to further clarify the effect of hymecromone on COVID-19 in a larger sample of clinical trials. Our findings highlight that inhibiting the synthesis of hyaluronan with specific drugs is a promising therapeutic strategy for COVID-19.

## Data Availability

In this study, the personal data of patients are sensitive and can't be shared in public. However, requests for data could be made to the Shanghai Public Health Clinical Center.

## Contributors

F.S., W.Y., and H.L. conceived and designed this study. S.Y., Y.L., F.Z., L.W., Z.S., M.L., Y.T., L. C., B.Z.,, Y.S. and F.S. did the follow-up investigation. S.Y., Y.L., F.Z, L.W., Q.L., D.R., T.Z., K.Z., P.X., and Z.Y. collected the data. S.Y., W.L., Q.L., and W.L. did the statistical analysis. S.Y. and F.Z. drafted the initial manuscript and all the authors revised the manuscript critically. H.L. and W.Y. had full access to the all data and took responsibility for the integrity and accuracy of the data in the study.

## Acknowledgments

This work was supported by the National Key R&D Program of China (2018YFC1005004), Major Special Projects of Basic Research of Shanghai Science and Technology Commission (18JC1411101), Shanghai Science and Technology Innovation Action Plan, Medical Innovation Research Special Project (20Z11900900). We thank Yue Yu for her editorial help and comments on the manuscript. We thank Ying Guo and Cuiyun Zhu for their help on patient data collection. We thank all the participants involved in this study. We also appreciate the assistance and support of Shanghai Public Health Clinical Center.

## Supplementary materials

**Supplementary Figure 1: Correlation between hyaluronan and subtype of lymphocytes cell counts in COVID-19 patients**

(A-C) The scatter plots showing CD4+ cell counts (A), CD8+ cell counts (B), and CD45+ cell counts (C) of COVID-19 patients classified by the 48.43 ng/ml of hyaluronan. Data was expressed by Mean ± SD. The significant difference was confirmed by Mann–Whitney test. *, *P* < 0.05; **, *P* < 0.01; ***, *P* < 0.001; ****, *P* < 0.0001; ns, not significant. (D-F) Counts of CD4+ cells (D), CD8+ cells (E), and CD45+ cells (F) upon admission plotted against hyaluronic acid, calculated by two-tailed Spearman’s correlation analysis.

**Supplementary Figure 2: hyaluronan and pulmonary lesions in severe COVID-19 patients**

(A-B) Scatter plot showing relation on the mass (A) and volume (B) of GGO against hyaluronic acid. (C-D) Scatter plot showing relation on the mass (C) and volume (D) of the consolidation region against hyaluronic acid. Mass and volume of pulmonary lesions were calculated via the automatic lung segmentation technology of AI based on CT images of severe COVID-19 patients. GGO was defined a range from -750 HU to -300 HU, and the consolidation region was defined from -300 HU to 50 HU. Two-tailed Spearman’s correlation analysis was performed to identify the relation of pulmonary lesions against hyaluronic acid.

**Supplementary Figure 3: hymecromone helps to decrease the CRP in COVID-19 patients**

Changes in lymphocytes (A), CRP (B), fibrinogen (C), and D-dimer (D) were calculated as the fold change of diverse clinical indicators per day in patients with CRP elevation. Data was presented by Mean ± SD. The significant difference was confirmed by the Mann–Whitney test. *, *P* < 0.05; **, *P* < 0.01; ***, *P* < 0.001; ****, *P* < 0.0001; ns, not significant.

**Supplementary Figure 4: hymecromone helps to decrease fibrinogen in COVID-19 patients**

Changes in lymphocytes (A), CRP (B), fibrinogen (C), and D-dimer (D) were calculated as the fold change of diverse clinical indicators per day in patients with fibrinogen elevation. Data was presented by Mean ± SD. The significant difference was confirmed by the Mann–Whitney test. *, *P* < 0.05; **, *P* < 0.01; ***, *P* < 0.001; ****, *P* < 0.0001; ns, not significant.

**Supplementary Figure 5: The effect of hymecromone on the elevation of D-dimer in COVID-19 patients**

Changes in lymphocytes (A), CRP (B), fibrinogen (C), and D-dimer (D) were calculated as the fold change of diverse clinical indicators per day in patients with D-dimer elevation. Data was presented by Mean ± SD. The significant difference was confirmed by the Mann–Whitney test. *, *P* < 0.05; **, *P* < 0.01; ***, *P* < 0.001; ****, *P* < 0.0001; ns, not significant.

**Supplementary Table 1: The primer sequences for RT-qPCR**.

