## Supplementary figures and images for "Hymecromone: A Clinical Prescription Hyaluronan Inhibitor for Efficiently Blocking COVID-19 Progression"

### Supplementary Figure 1

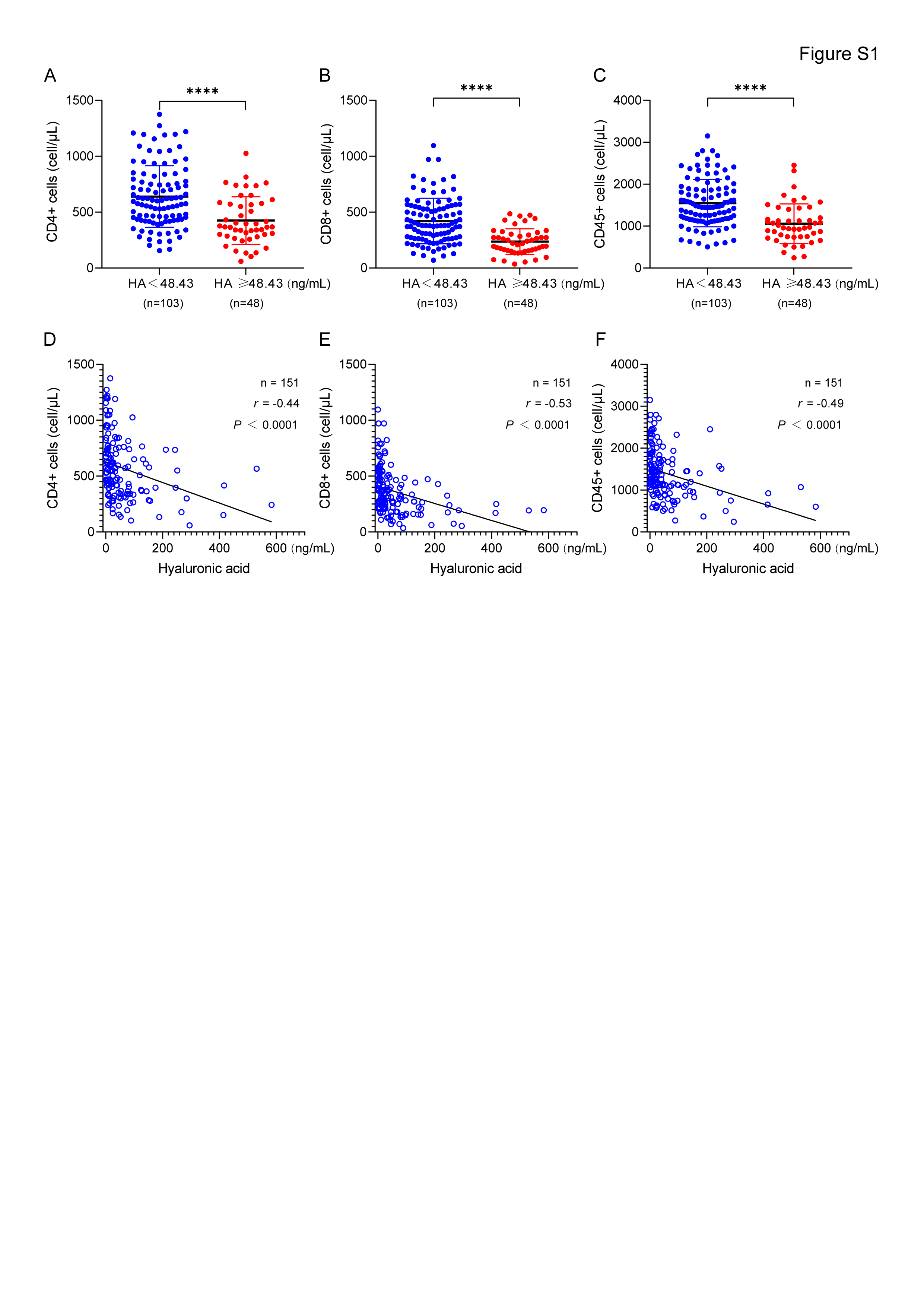

### Supplementary Figure 2

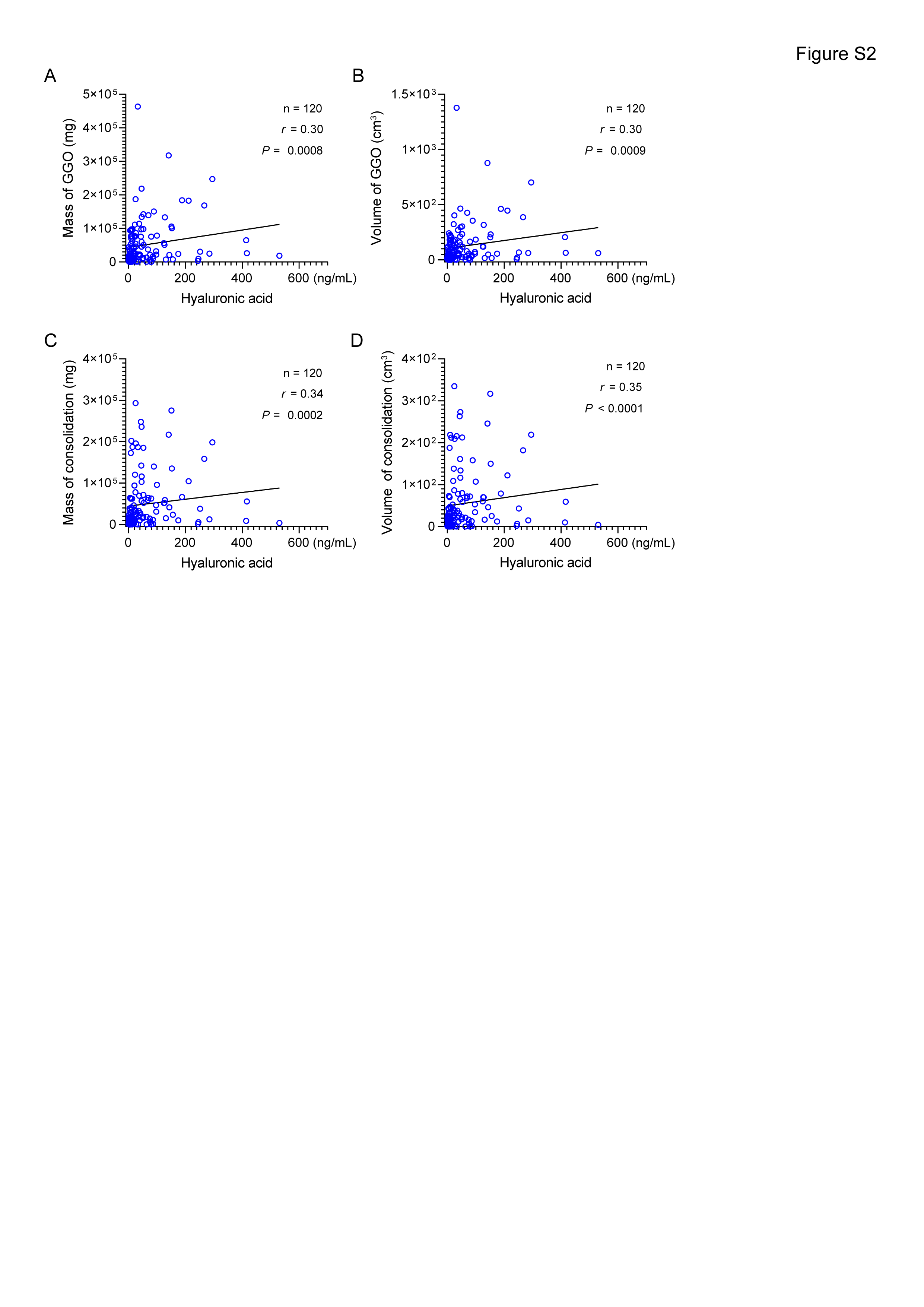

### Supplementary Figure 3

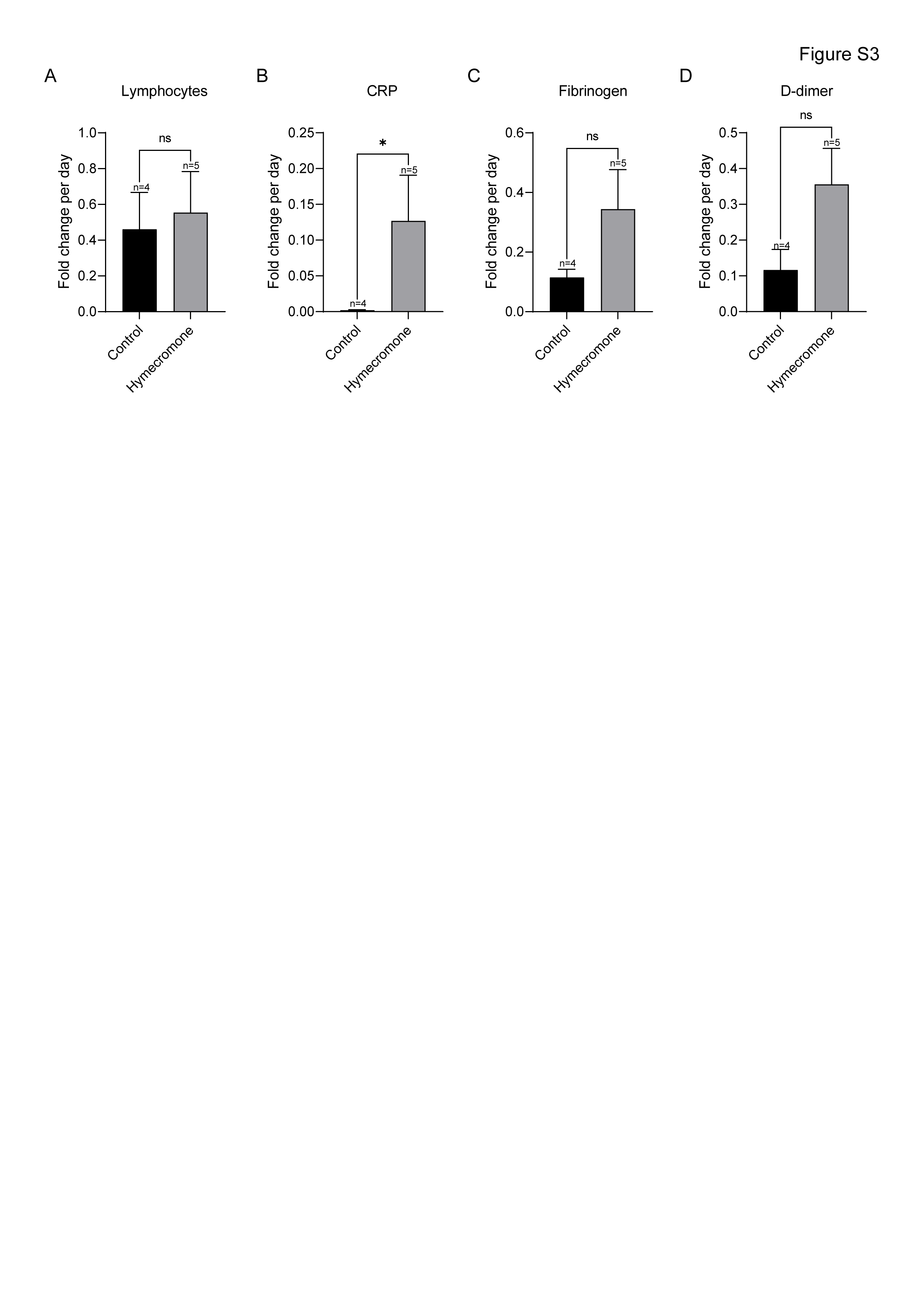

### Supplementary Figure 4

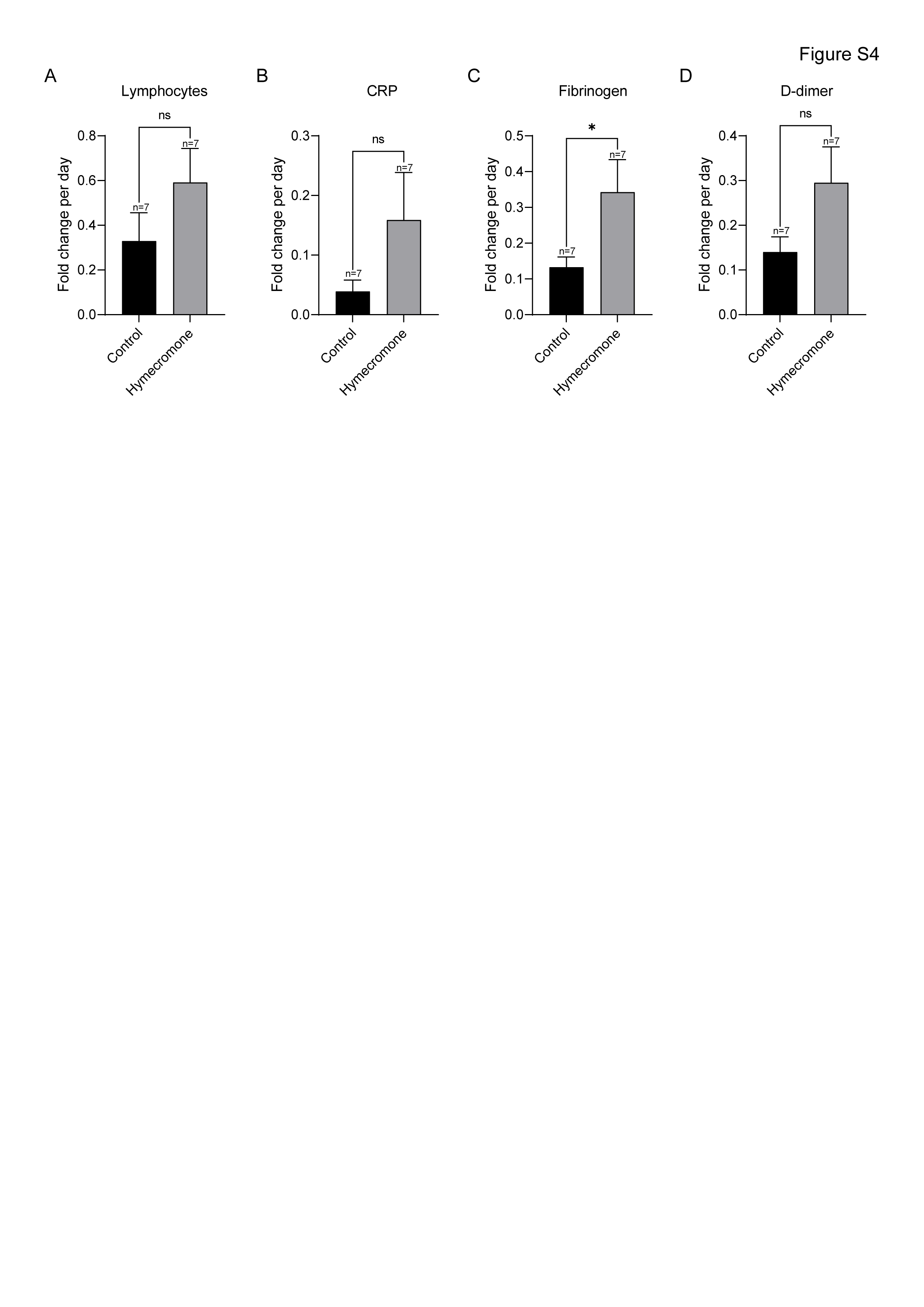

### Supplementary Figure 5

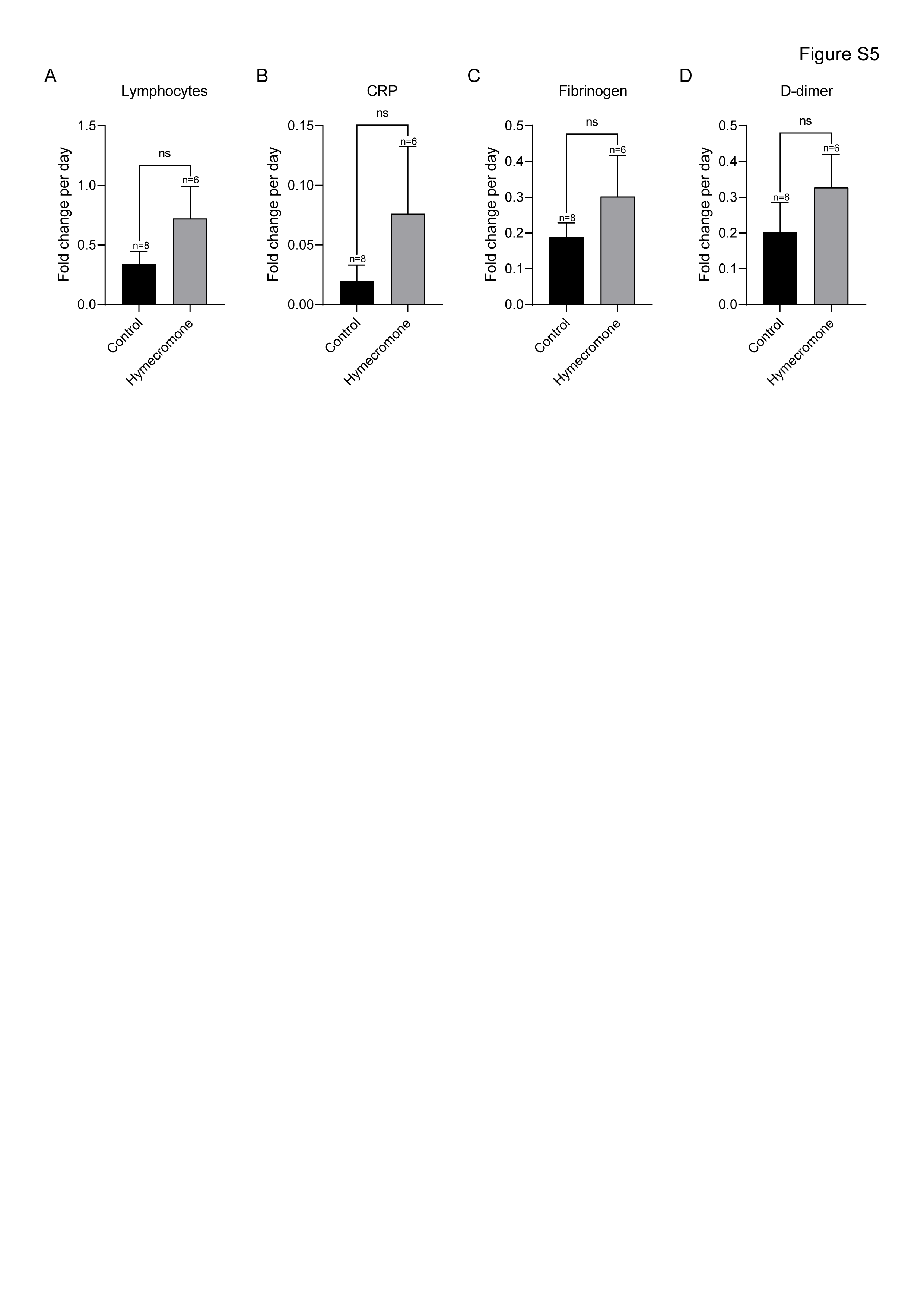
